# Molecular Spectrum and Chromatographic- Haematological Characterisation of Alpha Thalassemia Minor (-α/-α), and Alpha Silent Carriers (-α/-αα)

**DOI:** 10.1101/2023.05.02.23289372

**Authors:** Süheyl Uçucu

## Abstract

**Background and aims:** Alpha-thalassaemia is a group of disorders characterised by wide phenotypic variation caused by mutations in the α-globin genes (α1 and α2) of chromosome 16.

The aim of this study was to investigate the molecular profile of α-thalassemia variants and to compare and characterise the chromatographic behaviour and haematological properties of α-thalassemia minor (-α/-α, --/ααα) and α-silent carriers (-α/ααα) on HPLC.

**Materials & Methods:** A dataset of 200 individuals consisting of 42 alpha thalassemia minor (-α/-α, -- /ααα), 103 alpha silent carriers (-α/αα) and 55 normal participants from the Human Genetics Unit (HGU) of the Faculty of Medicine, Colombo, Srilanka was included. Blood samples from each patient were analysed by PCR for genotyping, haemogram and high performance liquid chromatography (HPLC) for characterisation. These data were then comparatively analysed using standard descriptive statistics.

**Results:** It was analysed in three sections as haematological, biochemical and molecular. Haematologically, alpha thalassaemia silent carrier was completely normal; alpha thalassaemia minor: decreased haemoglobin level; decreased MCV and MCH, normal RBC count. Alpha Thalassaemia Minor patients show a statistically significant difference from alpha silent carriers and normal population in terms of MCV, MCH, HGB, PCV, RDW.

Biochemically, alpha thalassaemia silent carriers were found to have normal alpha-globin chain production, alpha thalassaemia minor: low HBA2 as determined by HPLC. In terms of HPLC tests, similar results were observed between alpha thalassaemia minor patients and alpha silent carriers and between alpha thalassaemia minor patients and the normal group, with no statistical significance for all parameters except HBA2. Molecularly, the most common mutations in both variants are _α3.7 and _α4.2 mutations.

**Conclusion:** In summary, despite the haematological and biochemical differences between α-thalassaemia minor and normal individuals, both variants of alpha thalassaemia present a diagnostic conundrum, as CBC and HPLC results for individuals are comparable to normal humans. Although MCV, MCH, HGB, PCV and HBA2 levels differ between alpha thalassaemia minor carriers, alpha silent carriers and the normal group, conventional haemoglobin electrophoresis and haemogram alone have been found insufficient for the diagnosis of alpha thalassaemia. Therefore, although HPLC does not seem to be sufficient to distinguish between normal individuals and the two variants, the decrease in HBA2 levels is an important finding. Identifying a mutated alpha globin gene requires newer molecular diagnostic tests such as next generation sequencing (NGS) and quantitative PCR (qPCR). It should be noted that a given genotype can greatly alter the clinical manifestation by the presence of additional mutations, making the relationship between genotype and phenotype highly variable.

## Introduction

According to the World Health Organisation (WHO), more than one fifth of the world’s population carries one of the many different types of alpha (α) thalassaemia. According to WHO, it is estimated that approximately 15,000 babies worldwide will suffer from Hb-H Disease or Hydrops Fetalis of Haemoglobin Bart every year. It is most common in Mediterranean countries, Southeast Asia, Africa, the Middle East, South China and India, but is increasingly being reported in Northern Europe, North America and Australia due to migration waves. [1, 2].

Alpha thalassaemias (α-) are diseases with a wide phenotypic diversity caused by mutations in α-globin genes (α1 and α2) in the HBA gene on chromosome 16. The biochemical background of thalassaemia symptoms can manifest itself in various ways. Thalassaemias occur when there is an imbalance between the normal production of haemoglobin chain proteins (between alpha (HbA) and beta (HbB)) [3, 4]. HbA genes are located in the alpha globin cluster, while HbB genes are located in the beta globin cluster. Globin chains are regulated by globin genes in the alpha and beta globin gene clusters and by various components of the transcription machinery. As a result, abnormalities of these factors lead to a range of clinical features including microcytic hypochromic anaemia, ineffective erythropoiesis, growth retardation and psychomotor delay.

Alpha thalassaemia syndromes can present differently depending on the deletion of one or more alpha haemoglobin genes and can range from asymptomatic to hydrops foetalis. The number of deleted alpha genes will determine the type of this disease, which can be either deletion or point mutation for more severe cases, depending on how much is missing. According to the number of deleted alpha genes, it is referred to as asymptomatic (-α/-αα), carrier (-α/-α, - -/-α), HbH (- -/-α, -α/- -) and Hydrops fetalis (- -/-) [3, 5].. Hydrops fetalis is the form of alpha thalassaemia with the most severe clinical phenotype [6]. The defects are mainly directed towards deletion, so there is a large phenotypic variability. Depending on the genotype, symptoms of this disease may include anaemia, splenomegaly, liver dysfunction, bone deformities due to bone marrow failure and other complications of chronic haemolytic anaemia. Some clinical features are age-related; they may not be present at birth but are discovered later in life. Some symptoms may not appear immediately and some may not even appear until adulthood, but it is important for parents to be aware of them [7]. Both alpha thalassaemia minor (-α/-α, - -/-αα) and asymptomatic alpha silent carrier (-α/-α) have similar phenotypes and are benign [6]. However, a mutation that may accompany them may lead to a clinical phenotype. The degree of clinical phenotype severity associated with a given genotype can vary greatly with the presence of additional mutations, making the relationship between genotype and phenotype highly variable [6,8].

The aim of this study was to investigate the molecular spectrum of α-thalassemia variants, to compare and characterise the chromatographic behaviour and haematological properties of alpha silent carriers (-α/ααα) and alpha thalassemia minor (-α/-α, -- /ααα) on HPLC.

## Materials and Methods

This study used a database of 200 cases referred to the Human Genetics Unit (HGU) of Colombo Medical College, Colombo, Srilanka, between 2012 and 2020. The study included 103 alpha silent carriers (F=48, M=55), 55 normal participants (F=26, M=29), and 42 alpha thalassemia minor patients (F=20, M=22) diagnosed at the Human Genetics Unit of Colombo Medical College. Patients with iron deficiency anemia, beta-thalassemia, and other hemoglobinopathies were excluded from the patients diagnosed by genotyping in this study. The ratio of males to females was 51% to 49%.

Sociodemographic characteristics, hemogram, hemoglobin variant analysis, and DNA chain analysis data of the patients who were followed up at the Human Genetics Unit of Colombo Medical Faculty were obtained from the database of the relevant center. Informed consent forms were obtained from the patients.

Ethical approval for the study was obtained from PGIM Ethical Review Committee (Reference number: ERC/PGIM/2020/068) and the MSKU Ethical Review Committee (Reference number: 8428/11/2022), University of Colombo. The study was carried out under the Helsinki Declaration principles.

Serum samples were analyzed from EDTA-coated blood collected between 8:30 am and 3:00 pm. Erythrocyte index parameters were determined with an automated blood counting device Sysmex XN 1100 (Sysmex Diagnostic, Japan). Hemoglobin variant analysis was determined by high-pressure liquid chromo¬tography (HPLC) and Ion exchange method. Genotype data for the detection of Alpha-Talassemia mutations were analyzed by the Multiplex-Polymerase chain reaction (PCR) method.

Multiplex Polymerase Chain Reaction (PCR) is a powerful and efficient technique that allows the amplification of multiple DNA sequences in a single reaction. By amplifying multiple sequences simultaneously, this time- and the labor-saving process can significantly accelerate research and clinical diagnostics. To perform multiplex PCR, unique primers are required to lead to the amplification of specific regions of DNA, and methods to analyze individual amplification products from a mixture. Multiplex PCR has become a widely used and convenient screening test in both clinical and research laboratories and has the potential to enable the simultaneous detection of multiple agents that cause similar or identical clinical syndromes and/or share similar epidemiologic characteristics. In particular, Multiplex PCR is recognized as a highly effective method for the diagnosis of alpha-thalassemia.

### Statistical Analysis

The open-source statistical software Jamovi (version 2.2.5.0, Sydney, Australia) was employed to analyse the data. Descriptive statistics were calculated to provide an overview of the data, with frequency and percentage for categorical variables, and median and interquartile range (IQR) for numerical variables. The Shapiro-Wilk test was used to assess the conformity of continuous variables to a normal distribution. Meanwhile, Spearman’s correlation analysis was conducted to examine the relationship between numerical variables, as they did not meet the parametric test conditions. To gain a comprehensive understanding of the data and the hypotheses of the study, as the numerical variables did not meet this condition, the one-way ANOVA, a Kruskal-Wallis H Test (Non-parametric), and posthoc tests were used to compare the three independent groups Statistical significance was set at p<0.05.

## Results

In our sample group, there were 102 male and 98 female individuals. Of the total number of patients, 51% were male and 49% were female. The study included 103 alpha silent carriers (F=48, M=55), 55 normal participants (F=26, M=29) and 42 alpha thalassemia minor patients (F=20, M=22) diagnosed at the Human Genetics Unit of Colombo Medical School. The overall mean age was 13 years, mean (±Standard Deviation) male age was 14.76 (±19.21) and mean (±Standard Deviation) female age was 16.28 (±20.18).

In order to provide a detailed and informative comparison of the groups, it is possible to see the comprehensive hematological and biochemical profile analysis performed to compare Alpha Silent Carriers, Alpha Thalassemia Minors, and Healthy Individuals (Table 1).

**Table 1.** Unveiling the Haematological and Biochemical Profiles of Alpha Silent Carriers, Alpha Thalassemia Minors, and Healthy Individuals.

|  | Carrier status | Hb | PCV | RBC | MCV | MCH | MCHC | RDW | WBC | Neut | Lymph | PLT | HbA | HbA2 | HbF |
| --- | --- | --- | --- | --- | --- | --- | --- | --- | --- | --- | --- | --- | --- | --- | --- |
| 25th percentile (Q1) | alpha trait | 10.0 | 32.2 | 4.81 | 62.9 | 20.4 | 30.9 | 14.8 | 7.54 | 43.3 | 31.3 | 258 | 86.2 | 2.30 | 0.300 |
|  | normal | 10.7 | 33.3 | 4.74 | 65.5 | 20.7 | 31.4 | 13.6 | 8.5 | 41.4 | 36.5 | 260 | 85.8 | 2.45 | 0.200 |
|  | silent carrier | 10.9 | 33.7 | 4.70 | 67.5 | 21.5 | 31.4 | 13.4 | 7.60 | 42.0 | 37.0 | 268 | 86.1 | 2.50 | 0.300 |
| Median (Q2) | alpha trait | 10.8 | 33.1 | 5.21 | 68.5 | 20.9 | 31.7 | 16.0 | 9.92 | 48.2 | 41.5 | 354 | 87.3 | 2.47 | 0.561 |
|  | normal | 11.6 | 36.0 | 5.20 | 80.8 | 22.4 | 32.3 | 14.8 | 8.90 | 47.6 | 41.5 | 310 | 86.6 | 2.68 | 0.400 |
|  | silent carrier | 11.8 | 35.9 | 5.3 | 78.3 | 23.6 | 32.3 | 14.8 | 9.47 | 44.5 | 45.5 | 332 | 86.5 | 2.59 | 0.700 |
| 75th percentile (Q3) | alpha trait | 11.4 | 34.9 | 5.51 | 68.5 | 21.8 | 32.9 | 16.6 | 10.4 | 52.8 | 45.5 | 425 | 87.5 | 2.58 | 0.675 |
|  | normal | 13.4 | 39.1 | 5.45 | 80.8 | 26.6 | 33.5 | 15.9 | 10.6 | 52.0 | 47.2 | 382 | 87.5 | 2.80 | 0.735 |
|  | silent carrier | 12.7 | 37.2 | 5.43 | 78.3 | 25.6 | 33.0 | 16.0 | 11.4 | 52.0 | 49.0 | 396 | 87.3 | 2.70 | 0.769 |

**Note.** Hb: haemoglobin; RBC: red blood cell count; MCV: mean corpuscular volume; MCH: mean corpuscular volume; MCHC: mean corpuscular haemoglobin concentration, RDW: red distribution volume, WBC; Neut; Neutrophil, Lymph; lymphocyte, PLT; platelet, HbA2: haemoglobin A2; HbA0; haemoglobin A0, HbF; haemoglobin F

When we look at the molecular spectrum and percentage distribution of α-thalassemia minor (-α/-α, -- /αααα) and α-thalassemia silent carrier (-α/ααα) mutations, it is possible to see the -α3.7 mutation with the highest rate (Table 2).

**Table 2.** Molecular spectrum of α-thalassemia minor (-α/-α, -- /ααα) and α-thalassemia silent carrier (-α/ααα) mutations

| Phenotype | Genotype | Percentage (%) |
| --- | --- | --- |
| $\alpha$ -thalassemia trait | $-\alpha 3.7/-\alpha 3.7$ | 28% |
| | $-\alpha 3.7/-\alpha 4.2$ | 3.2% |
| | $-\alpha 4.2/-\alpha 4.2$ | 1.6% |
| $\alpha$ -thalassemia silent carrier | $aa/-\alpha 3.7$ | 56% |
| | $aa/-\alpha 4.2$ | 9.6% |
| Other* | -HbH | 1.6% |

Note. Excluded\*

When we look at Figure 1, it is possible to visually examine the numerical data and the variability of the data by displaying the distribution of the parameters between alpha thalassaemia minor patients, alpha silent carriers and the normal population, among which we found the highest statistical difference (Table 1).

**Figure 1.**
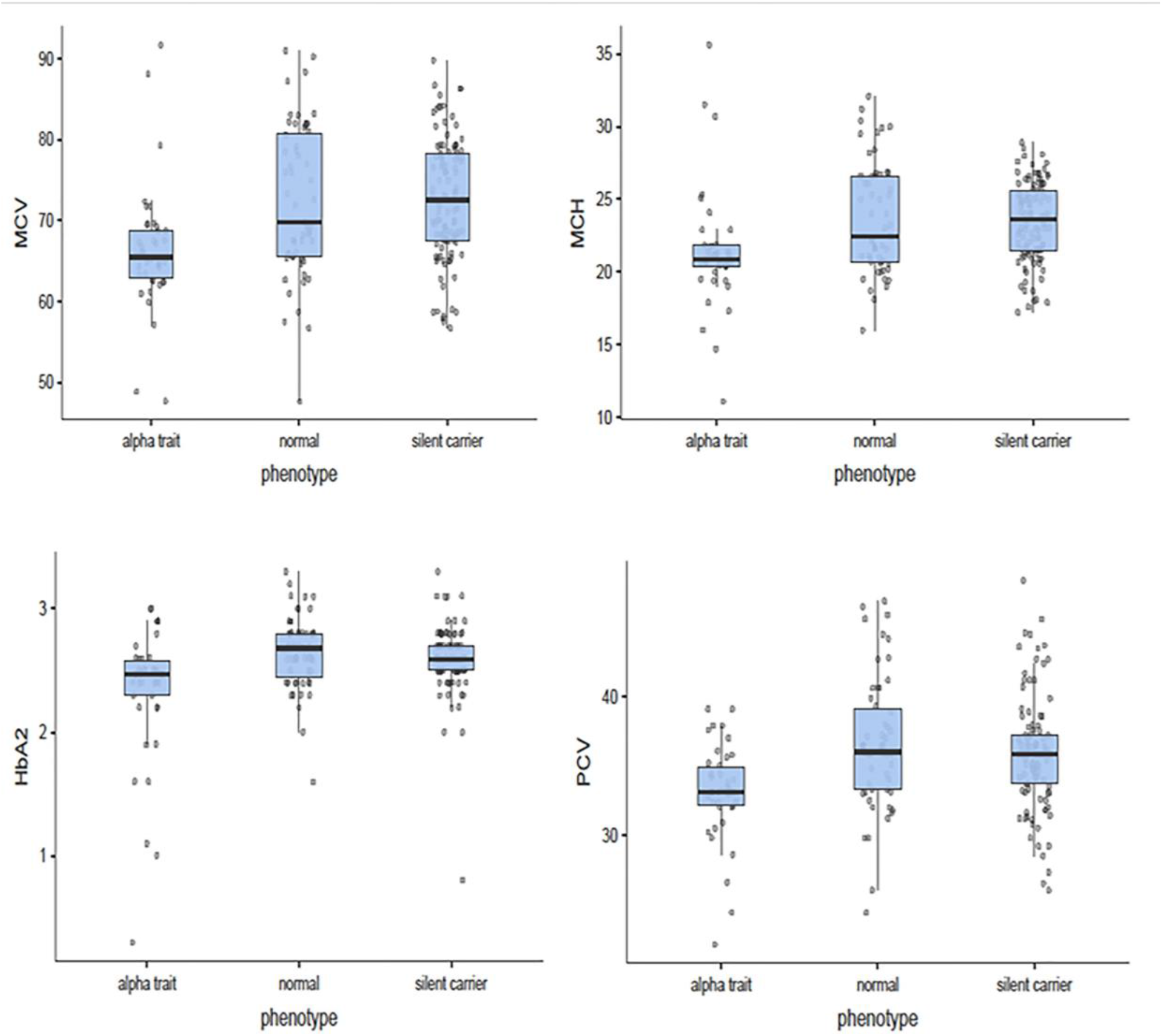
An Exploration of Box Plots of α-Thalassemia Minor (-α/-α, --/ααα), α-Thalassemia Silent Carrier (-α/αα) and Normal Groups

Table 3 shows that there is a statistical difference between the groups in terms of MCV, MCH, HGB, PCV, RDW and HbA2. However, we need post-hoc tests (presented in Table 4) to understand between which groups.

**Table 3.** Kruskal Wallis H Test (Non-parametric)

| Parameters | $\chi^2$ | df | p | $\epsilon^2$ |
| --- | --- | --- | --- | --- |
| Hb | 16.83 | 2 | < .001 | **0.08371** |
| PCV | 18.29 | 2 | < .001 | **0.09098** |
| RBC | 2.20 | 2 | 0.333 | 0.01095 |
| MCV | 22.03 | 2 | < .001 | **0.10960** |
| MCH | 18.57 | 2 | < .001 | **0.09287** |
| RDW | 16.03 | 2 | < .001 | **0.07976** |
| WBC | 1.19 | 2 | 0.551 | 0.00593 |
| Neut | 5.14 | 2 | 0.077 | 0.02556 |
| Lymph | 4.67 | 2 | 0.097 | 0.02321 |
| PLT | 2.53 | 2 | 0.283 | 0.01257 |
| HbA | 3.60 | 2 | 0.165 | 0.01793 |
| HbA2 | 18.82 | 2 | < .001 | **0.09364** |
| HbF | 3.94 | 2 | 0.140 | 0.01958 |

Note. $\chi^2$ : Asymptotic significance, df: Degrees of freedom, $\epsilon^2$ : epsilon, p: Alpha significance $p < 0.05^*$ , Hb: haemoglobin; RBC: red blood cell count; MCV: mean corpuscular volume; MCH: mean corpuscular volume; MCHC: mean corpuscular haemoglobin concentration, PCV: packed cell volume, RDW: red distribution volume, WBC: neutrophil, Lymph: lymphocyte, PLT: platelet, HbA2: haemoglobin A2, HbA0: haemoglobin A0, HbF: haemoglobin F

**Table 4.**
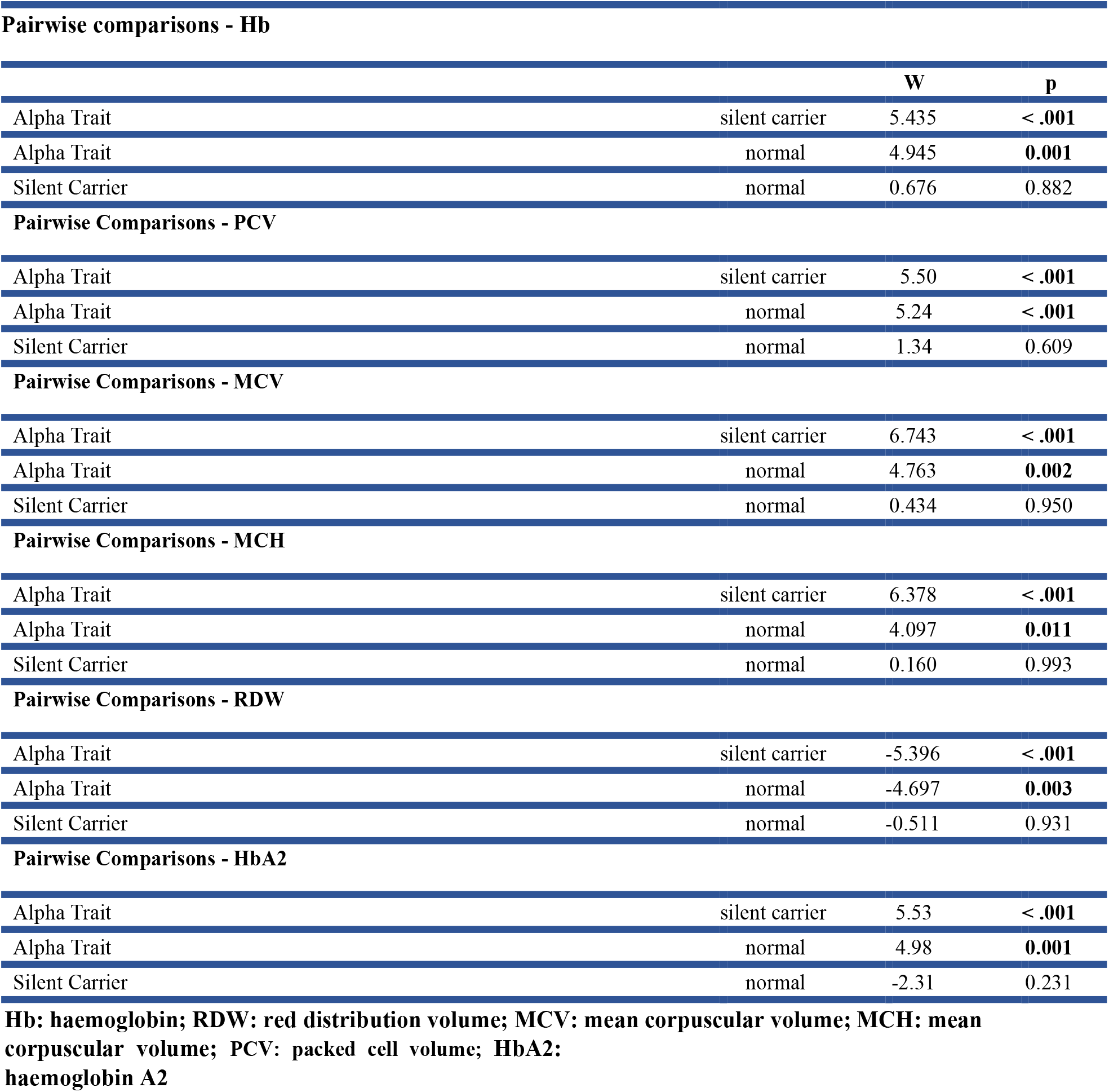
Exploring the Differences: A Comparative Analysis of α-Thalassemia Minor (-α/-α, -/ααα), α-Thalassemia Silent Carrier (-α/-αα) and Normal Groups.

Table 4 shows that there is a statistical disparity between alpha thalassaemia minor patients, alpha silent carriers and the normal population in terms of MCV, MCH, HGB, PCV, RDW and HbA2. No statistical differences were detected between alpha silent carriers and the normal group. It is possible to see that the HPLC tests gave similar results between alpha minor and alpha silent carrier and between alpha thalassemia minor and the normal group with statistically significant differences, whereas between alpha silent carrier and the normal group all parameters were of no statistical significance (Tables 3, 4).

## Discussion

In this study, we investigated the chromatographic and haematological profiles of alpha thalassaemia minor (-α/-α, - -/ααα) and alpha silent carriers (-α/αα) and the molecular spectrum of α-thalassaemia mutations.

The binary structure of human haemoglobin is complex and consists of two genes, each with two separate chromosomes (chromosome 16 for alpha globins and chromosome 11 for beta globins). The embryonic gene, two fetal/adult genes, one unit of fetal G and one unit of epsilon gene are the five such functional genes in this cluster. Located on chromosome 16, the alpha-globin genes (1 and 2) are the source of the phenotypic diversity of alpha-thalassaemias.

The key to the diagnosis of alpha thalassaemia is to perform a comprehensive blood count. This is the first step in the diagnostic process and a review of the person’s medical history should be followed by examination of serum ferritin levels, MCV and MCH levels and other indicators of iron deficiency anaemia. If these tests exclude iron deficiency anaemia, methods such as HPLC, capillary electrophoresis or haemoglobin electrophoresis are sought to test for levels of haemoglobin variants. These methods measure the amount of haemoglobin present and can distinguish between the different types of haemoglobin present in the blood sample [3,6]. For the silent carrier of alpha thalassaemia, haemoglobin electrophoresis usually gives a normal result. In this case, the only way to accurately determine whether one of the alpha globin genes is mutated is to perform DNA analysis by molecular testing. If iron deficiency, beta thalassaemia carriage and anaemia of chronic disease are ruled out by routine blood tests in these individuals, it is safe to conclude that they have alpha-thalassaemia by molecular testing [3,6].

According to our findings, there are interesting differences between α-thalassemia minor (-α/α,-- /ααα) and α-thalassemia silent carriers (-α/αα) and normal group in terms of chromatographic and haematological characteristics (MCV, MCH, HGB, HBA2 and PCV). The primary difference between silent carriers and minor thalassaemia is the amount of haemoglobin and haemoglobin A2 produced by the body. While haemoglobin A2 levels were normal in silent carrier individuals, individuals with minor thalassaemia had microcytic hypochromic findings and a significant HBA2 decrease at the sample level (P value <0.001). However, there was no significant difference in HbA2 between alpha silent carriers and the group of normal individuals. Similarly, HPLC analyses from all three groups gave similar and statistically insignificant results except for HbA2. According to studies, individuals with alpha-thalassaemia silent carriers typically have normal haemoglobin levels and no other haematological abnormalities, while microcytic hypochromic findings and low HbA2 levels have been reported in alpha-thalassaemia minors. [3,5,6]. In our study, in agreement with these studies, silent carriers typically have a normal haemoglobin level on CBC and normal alpha-globin levels on HPLC, whereas alpha-thalassemia minors usually have low alpha-globin levels on CBC and HPLC. The possible reason for this may be that only one alpha-globin gene is mutated in α-thalassemia silent carriers, so there is still sufficient functional haemoglobin to maintain normal levels. However, individuals with alphathalassemia minor often have reduced haemoglobin levels due to the presence of two mutated alpha globin genes. Decreased haemoglobin levels can lead to other haematological abnormalities such as anaemia or microcytic anaemia [3,6]. This may be a possible reason for the differences in MCV and MCH in individuals with alpha thalassaemia minor that we observed in our findings. It is important to separate and measure haemoglobin fractions together with the blood count to ensure that any potential abnormality in the normocytic range is not overlooked. However, in multi-ethnic populations, the use of MCV and MCH in the differential diagnosis is not recommended [2]. Although RDW is higher in alpha thalassaemia minors, we believe that it is not a reliable discrimination marker since the values are comparable with normal individuals and the reference range [9].

In addition, alpha-thalassaemia minor can cause mild symptoms such as fatigue or pale skin, while the silent feature of alpha-thalassaemia appears to be a haematological abnormality and not a clinical symptom. It appears to be an inherited blood disorder caused by a genetic defect that reduces the production of alpha-globin, one of the main proteins involved in haemoglobin formation. Individuals with the silent feature of alpha-thalassaemia usually have no clinical symptoms and are only identified by laboratory tests.

However, it should be noted that although HbA2 appears to be statistically different between α-thalassemia minor and normal individuals at the sample size, at the individual level, both alpha-thalassemia variants are a problem for differential diagnosis, as HPLC results appear to be similar to those in normal individuals (α-thalassemia minor HbA2: 2.47%, α-thalassemia silent HbA2: 2.59%, normal HbA2: 2.68%). Therefore, HPLC is an important finding, although it does not seem to be sufficient to distinguish between both normal individuals and the two variants. Similarly, it should be kept in mind that parameters such as MCV, MCH, HBG, which seem to be important differential findings, can be affected by many different conditions such as megaloblastic anaemia, iron deficiency anaemia, gammopathies, hypothyroidism, myelodysplastic syndrome [6].

From a molecular perspective, it is possible to gain a deeper understanding of the genetic dynamics of alpha-thalassaemias by analysing them at the molecular level to gain insights that can help us develop better strategies [3].Without molecular analysis it is not possible to fully understand the molecular dynamics of alpha-thalassaemia and thus reduce its prevalence [3,10].Unfortunately, the cost of molecular testing can be a significant barrier to screening in developing countries; it may therefore prevent a full understanding of the prevalence of alpha-thalassaemia in the population, its regional distribution, the coexistence of alpha and beta-thalassaemia mutations and the clinical phenotype it will produce. The degree of severity of the clinical phenotype associated with a given genotype can vary greatly with the presence of additional mutations, making the relationship between genotype and phenotype highly variable [8].Higgs et al. discussed the molecular basis of alpha-thalassemia, how it is caused by deletions in the alpha-globin gene cluster, and the genotype-phenotype relationship. The authors reported that in some cases it is difficult to associate phenotype with genotype (genetic makeup) due to the heterogeneous nature of the underlying abnormalities. However, individuals with alpha-thalassemia may have reduced red blood cell and hemoglobin levels, which can be detected by a complete blood count test [12,13].

According to studies, the most common alpha-thalassaemia variant in the world is α3.7 (-α/ααα). According to the papers, The prevalence of alpha thalassemia mutation 3.7 is high in a number of places, including the Netherlands (58.2%), Sicily (46.9%), Spain (52.4%), Malaysia (45.9%), northern Thailand (10.7%), and Brazil (10.7%). In East Asia, it is rare. Alpha thalassemia variants -3.7 (60%) and -4.2 are the most prevalent in Iran, and more than 70 non-deletional mutations have also been found. Iran (1.8-4.8%), Sicily (4.5%), Greece (12%), the Netherlands (0.9%), and Northern Cyprus (7.8%) all have varied rates of -20.5 double gene deletion [14-17].Studies on alpha thalassemia in Turkey identified 14 distinct variants, with the -3.7 deletion being the most prevalent [18-21].Studies conducted in Sri Lanka revealed that α3.7 (-α/ααα) and 4.2 kb mutations are the most common alpha-thalassemia gene defects [11]. In the study group we used, the mutation responsible for 56% of α-thalassemia silent carriers was αα/_α3.7, while the mutation responsible for 28% of α-thalassemia minors was _α3.7/_α3.7. This is consistent with the global trend of α3.7 (-α/ααα) being the most common mutation in α-thalassaemia [11,21].

In conclusion, the haematological, biochemical and molecular examination of the two alpha thalassaemia variants shows that despite differences in MCV, MCH, HGB, PCV and HBA2 levels, conventional haemoglobin electrophoresis, HPLC and haemogram alone are insufficient for the diagnosis of alpha thalassaemia. From a molecular point of view, although alpha thalassaemia minor and silent carriage may appear innocent, the clinical manifestation of a given genotype may vary greatly with the presence of additional mutations, making the relationship between genotype and phenotype highly variable.

## Conclusion

In summary, despite the haematological and biochemical differences between α-thalassaemia minor and normal individuals, both variants of alpha thalassaemia present a diagnostic conundrum, as CBC and HPLC results for individuals are comparable to normal humans. Although MCV, MCH, HGB, PCV and HBA2 levels differ between alpha thalassaemia minor carriers, alpha silent carriers and the normal group, conventional haemoglobin electrophoresis and haemogram alone have been found insufficient for the diagnosis of alpha thalassaemia. Therefore, although HPLC does not seem to be sufficient to distinguish between both normal individuals and the two variants, the decrease in HBA2 levels is an important finding.

As a result, alpha thalassaemia is often not diagnosed by normal haemoglobin electrophoresis and haemogram, and identifying a mutated alpha globin gene requires newer molecular diagnostic tests such as next generation sequencing (NGS) and quantitative PCR (qPCR) [12,13]. It should be noted that a given genotype may greatly modify the clinical manifestation by the presence of additional mutations, making the relationship between genotype and phenotype highly variable. Investigation of the topic using data from a larger number of participants would be extremely useful to confirm and improve our conclusions.

## Data Availability

All data produced in the present work are contained in the manuscript

## Ethical standards

‘ All procedures performed in studies involving human participants were in accordance with the ethical standards of the institutional and/or national research committee and with the 1964 Helsinki Declaration and its later amendments or comparable ethical standards. ’

## Funding

None funding. The funders had no role in study design, data collection, and analysis, decision to publish, or preparation of the manuscript.

## Conflict of Interest

The authors declare no conflict of interest.

## Notes

### Competing Interest Statement

There is not conflict of interest.

### Funding Statement

This study did not receive any funding

### Author Declarations

Ethical approval for the study was obtained from PGIM Ethical Review Committee (Reference number: ERC/PGIM/2020/068) and the MSKU Ethical Review Committee (Reference number: 8428/11/2022), University of Colombo.

